# Emergency Department Discharge Center Program Evaluation from a “Learning Organization” lens: Methods, Lessons Learned, and Future Directions

**DOI:** 10.1101/2024.07.30.24310873

**Authors:** Bibi S. Razack, Naya Mahabir, Lisa Iyeke, Lindsay Jordan, Roland Hope, Emily Diaz, Lyze Barcia, Diana Fuzailov, Helena Willis, Marina Gizzi-Murphy, Frederick Davis, Adam Berman, Mark Richman, Nancy Kwon

## Abstract

Our ED’s Discharge Center (EDDC) facilitates appointments and paper-based social determinants of health (SDoH) screening. No criteria guide EDDC utilization. The ED’s provider-administrator-run, patient-satisfying follow-up call program contacts only ∼25% of discharges. We describe Learning Organization-principle-guided evaluation of EDDC efficiency, aiming to create EDDC time to expand the follow-up program.

We reviewed appointment-making, SDoH-screening, and follow-up program data. We surveyed patients to determine whether adopting SHOUT tool criteria (no home, no primary care physician, or insurance) might yield more-judicious EDDC utilization.

EDDC staff’s 20 minutes/patient yielded fewer ED returns and admissions. Most patients improved post-discharge and made appointments themselves; 6% met SHOUT criteria for EDDC assistance; 4.5% would benefit from SDoH screening.

Adopting SHOUT criteria would create significant time for EDDC-staffed follow-up program expansion. QR-code-accessible SDoH surveys would screen thousands more patients, minimizing EDDC staff involvement, saving 95% of the effort while retaining 100% of the benefit.

## Introduction

In July 2020, the Northwell Health Long Island Jewish (LIJ) Medical Center Emergency Department instituted an Emergency Department Discharge Center (EDDC). This innovative program aims to: 1) improve patient follow-up after ED discharge by having Care Coordinators facilitate patient follow-up appointments prior to discharge; 2) increase the screening for social determinants of health (SDoH) and referral to community-based organization (CBO) resources, as needed, and 3) decrease health disparities in vulnerable patient populations.

Timely follow-up after discharge from the ED improves patient-centered outcomes such as 30-day admissions.^1^ However, after discharge, many patients have difficulty securing and attending appointments due to challenges including lack of insurance, misunderstanding discharge instructions, and overlooking the importance of follow-up care.^2,3^ Many ED patients lack a primary care physician (PCP) or specialist, making it difficult to access timely healthcare.^4^ Even when patients have a PCP or specialist, these providers are often unaware their patients visited an ED. Patients with a PCP or specialist may still have difficulty arranging their own follow-up on account of lack of appointment availability (a situation unlikely to improve based on projections of physician supply and demand),^5^ inadequate insurance, poor health literacy, low self-efficacy,, or non-adherence to their treatment plan.^6,7^ Patients who leave the ED with only a phone number rather than a specific appointment are less likely to attend a discharge appointment within 10 days.^8^

The EDDC has been described elsewhere.^9^ Briefly, LIJ is a 583-bed hospital serving a racially and socioeconomically diverse population that includes many uninsured or underinsured patients. The adult ED sees ∼100,000 patients and discharges ∼75,000 patients annually. The typical discharge process for “non-EDDC” patients consists of providers explaining the patient’s confirmed or suspected condition and return precautions; providing lab and imaging results; and giving a list of relevant follow-up specialists’ or PCPs’ names and contact information. Patients are responsible for reaching out to those physicians or to physicians with whom they have an existing relationship.

The EDDC operates Monday-Friday, 7 AM-5 PM, and occasionally Saturdays. It is housed in a quiet area within the ED. The center is staffed by four Care Coordinators. There are no criteria/guidelines or restrictions on the characteristics of patients (eg, age, insurance, diagnoses) who can be referred to the EDDC. Care Coordinators obtain from the patient or provider the following patient information to make an appointment: 1) specialty and/or PCP follow-up care required; 2) diagnosis; 3) follow-up time frame; 4) health insurance; and 5) contact information. EDDC staff work with the patient to select a preferred appointment date, time, and location. Referral appointments are limited to Northwell locations.

During the EDDC pilot year, 3,616 patients were managed by the EDDC (6.4% of all discharged patients). EDDC staffing has since increased and is on pace to assist 12,000 patients this year (16% of discharges). In unadjusted analysis, EDDC patients were slightly less likely to return to the ED within 72 hours (5.3% vs. 6.5%; p = 0.0044; NNT = 83) or be hospitalized within 30 days (3.4% vs. 4.2%; p = 0.033; NNT = 143) compared to non-EDDC discharged patients. These effects were driven by assisting profoundly vulnerable patients. Among uninsured patients, 5.9% of EDDC patients had a 72-hour return vs. 2.4% of non-EDDC patients (p = 0.003), and 4.3% had a 30-day admission vs. 1.0% of non-EDDC patients (p = <0.0001). Of patients 65 and older, 5.7% of EDDC patients had a 30-day admission vs. 9.2% of non-EDDC patients (p = 0.005). Of patients without a PCP, 6.0% of EDDC patients vs. 7.4% of non-EDDC patients (p = 0.0647) had a 72-hour return.

Providers vary greatly in whom they refer to the EDDC, including patients who: 1) need a PCP or specialist, 2) require 1-2 week follow-up, 3) have poor health literacy, 4) don’t speak English, 5) already had difficulty making an appointment, and 6) would be admitted if not for a timely appointment. EDDC staff also screen patients for SDoH using a printed form. Respondents indicating a social services need (eg, housing, transportation) are given to an ED secretary, who manually enters patient identifiers, contact information, and particular social service needs data into a commercially-available social services portal (NOWPOW®), which then texts or emails patient names and contact information of local non-profit community-based organizations (CBOs). That secretary then enters largely the same information into a separate ED-created, HIPAA-compliant REDCap® database. During the pilot year, EDDC staff screened 565 patients (1% of discharges) for SDoH and are on pace to screen 5,000 patients this year (7% of discharges).

The ED has a separate initiative to create a post-discharge safety net and improve patient satisfaction. For the past 2 years, the ED has operated a follow-up call program staffed by physician administrators and physician assistant administrators. A random selection of patients is called back by ED physicians (60%), nurses (23%), or administrative staff (17%). Patients are queried about the course of their condition, whether they have follow-up appointments, and if they have any questions related to their visit. Data on approximately 4,000 calls made within 1 week of discharge indicated the majority of patients (∼77%) felt better or the same (∼21%). This program is associated with higher satisfaction scores; but, only a random 25% are called, due to volume.

EDDC leadership hypothesized that adopting criteria for providers to refer to the EDDC and simplifying the SDoH screening process would improve the efficiency of both processes, as measured by fewer patients needing to be “touched” by the EDDC to result in a prevented 72-hour return or 30-day admission (ie, a lower NNT for 72-hour returns and 30-day readmissions). The improved efficiency could be used to expand the follow-up call program by delegating EDDC staff to perform such calls. This article describes the process by which we utilized “Learning Organization” principles to evaluate the EDDC for opportunities for improved efficiency.

## Methods

A “Learning Organization” is one which follows Peter Senge‘s 5 principles (systems thinking, personal mastery, mental models, shared vision, and team learning) to “facilitate the learning of its members and continuously transform itself.”^10^ To evaluate EDDC processes (“systems thinking”), the overriding Learning Organization principle we adopted was “integrate learning into the business process.” We established “team learning” by engaging EDDC staff and ED leadership (“leadership commitment”), thereby “promoting ownership at every level.” We shadowed EDDC staff and reviewed data for 3,616 patients receiving appointment assistance and 342 offered SDoH screening over a 1-year period, and 4,877 followed-up by phone over a 2-year period. Over the course of one month, we randomly surveyed 50 patients for how many would benefit from EDDC assistance if we used validated SHOUT^11^ tool criteria for discharge assistance, and how many had SDoH social service issues **plus** the ability to follow through with contacting CBOs. The SHOUT tool screens for risk factors for “discharge failure” (short-term return to the ED); such risk factors are weighted (eg, male = 2 points, lack of PCP = 21 points). For ease of patient, provider, and staff use (ie, minimizing the number of screening items), our survey asked patients whether they met any of the highest-weighted risks (homelessness, lack of PCP, uninsured).

## Results

### Key observations

EDDC staff require an average of 20 minutes per patient they assist to obtain an appointment; this time comprises interviewing the patient, reaching out to clinics, and entering data. EDDC staff don’t make appointments. Rather, they act as liaisons, calling and/or emailing physicians’ offices to facilitate appointment scheduling on behalf of the patient while verifying the clinic accepts the patient’s insurance.

Regarding SDoH screening, of all patients offered the screening survey, only 31% completed it; 20% identified a need (65% of those who completed it); and only 5% completed it, identified a need, and followed-up on their own after receiving texted or emailed CBO names and contact information. The ED secretary manually enters data for all patients who identify a social service need into the ED-created REDCap® database, despite low rates of patients availing themselves of CBOs to which they are referred (that is, only one-fourth of those referred to a CBO follow through with contacting that CBO). We discovered the ED-created REDCap® database is largely superfluous, as it is never queried and its data is not used for planning or evaluation; for example, the ED has no plans to hire a social worker to address needs entered into REDCap®.

Data from the follow-up call program found 98% of patients stated the problem for which they attended the ED had either improved or was the same; 50% had gotten a 1-week follow-up appointment without EDDC assistance; only 10% had clinical questions.

The random survey revealed only 6% of patients meet SHOUT tool criteria for EDDC appointment assistance services (currently, 16% are referred to the EDDC), and 4.5% need and would contact CBOs for SDoH services.

### Lessons learned

1. EDDC staff help facilitate appointments by contacting clinics, verifying insurance, and addressing barriers to follow-up. While the patient might not leave with a time and date for the follow up appointment, they will be in the system for the physician’s office to call back to help arrange.
2. The EDDC makes a notable, but small, difference in 72-hour returns (NNT = 83) and 30-day admissions (NNT = 143). While some patients might be able to call on their own for an appointment, having EDDC staff who know which insurances are taken and who direct the initial call facilitates the process. The absence of criteria for referral to the EDDC likely contributes to these high NNTs, as many patients who won’t benefit from the EDDC’s appointment-facilitating service (because they have a PCP or specialist or can make their own appointments) are referred to the EDDC, inflating the NNT.
3. Only ∼5% of patients offered the SDOH survey did all of these three: 1) agreed to participate in the survey, 2) identified a social need, and 3) were willing and able to follow up with community-based organizations. With such low rates, in order to assist a substantial number of patients, the screening will have to become more widely distributed and electronic, rather than paper-based.
4. EDDC staff are a flexible resource who can be deployed for:
  1. Assistance making appointments.
  2. SDoH screening and connection to resources
  3. Making follow-up calls

### Future directions

The Learning Organization evaluation process found that adopting SHOUT criteria for screening to the EDDC for appointment assistance would improve efficiency, reducing referrals by ∼⅔ (from 16% to 6% = 7,500 fewer low-yield referrals). This would free 2,500 hours for EDDC to staff the call-back program, serving as a first-line screen, with the minority of patients with clinical questions referred to a provider.

Because only a few percent of patients complete the SDoH survey, identify a social need, and are willing/able to contact a CBO, the best way to find patients who might benefit from SDoH screening would be to screen many more than only the currently screened <10% of discharged patients. SDoH screening could be expanded by using an electronic screening questionnaire by which the patient can self-identify as meeting three criteria indicating they would benefit from EDDC assistance (willingness to complete the survey, having a social need, and willingness/ability to contact a CBO) **before** EDDC staff are utilized.

With this in mind, the group decided to:

1. Create one brief, QR code-accessible electronic tool incorporating SHOUT criteria and SDoH screening. Patients with any highly-weighted SHOUT risk (homelessness, no PCP, lack of uninsurance) will be eligible for EDDC assistance. The SDoH portion will utilize branching logic; patients will first be asked if they are willing and able to contact CBOs on their own. Those unable will not be asked further questions. This process should expand SDoH screening. Previously, EDDC staff distributed a paper-based survey to only 1% of discharged patients. Patients unwilling to complete the QR code survey, or who do not identify any social needs, or who are unwilling or unable to follow through on their own will never come to the EDDC staff’s attention. Only patients who complete the survey, identify a social need, and are willing and able to follow through on their own would occupy EDDC resources, thus saving 95% of the effort while retaining 100% of the benefit.
2. Eliminate redundant data entry.

What other organizations’ ED leadership can learn from our “Learning Organization” process and discoveries:

1. Overtly state “leadership commitment” to the process
2. Engage all levels of staff through regular meetings to “integrate learning into the business process” and create “team learning.”
3. Don’t focus on the performance of any particular individual. Instead, adopt a “systems thinking” approach.
4. Listen to front-line thinkers, thereby “promoting ownership at every level.”
5. Use open-ended questions, asked in a spirit of inquiry, not authority or punishment.
6. Dig deep: utilize techniques such as “5 Whys” to uncover underlying issues.
7. Collect data, but only what will be useful for planning or evaluation.
8. Review data and stories.
9. Change operations based on data.

## Conclusion

The “Learning Organization” approach to the EDDC program evaluation revealed opportunities to improve effectiveness and efficiency. Adopting SHOUT tool criteria for who can be referred to the EDDC, and an upfront electronic questionnaire that includes not only the need for social services, but also the interest and ability to carry through with contacting community-based organizations, may improve the return on investment of engaging patients and save thousands of hours of EDDC staff time per year. This time can be used to expand the ED’s follow-up call program while freeing up valuable physician administrator and physician assistant administrator time. As one of the issues patients called in follow-up may describe is difficulty obtaining appointments, the EDDC staff will be ideally suited to assist with this issue. EDDC staff can refer patients with clinical questions to an available physician administrator and physician assistant administrator.

## Data Availability

All data produced in the present study are available upon reasonable request to the authors.

## Notes

### Competing Interest Statement

The authors have declared no competing interest.

### Funding Statement

This study did not receive any funding.

### Author Declarations

This study was deemed exempt by Northwell Health's Human Subject Protection Program - Institutional Review Board (IRB) as a quality improvement project.

## References

1. Misky GJ, Wald HL, Coleman EA. Post-hospitalization transitions: Examining the effects of timing of primary care provider follow-up. J Hosp Med. 2010 Sep;5(7):392–7. doi: 10.1002/jhm.666. PMID: 20578046.

2. Mahajan M, Hogewoning JA, Zewald JJA, Kerkmeer M, Feitsma M, van Rijssel DA. The impact of teach-back on patient recall and understanding of discharge information in the emergency department: the Emergency Teach-Back (EM-TeBa) study. Int J Emerg Med. 2020 Sep 24;13(1):49. doi: 10.1186/s12245-020-00306-9. PMID: 32972361; PMCID: PMC7513274.

3. Mahajan M, Hogewoning JA, Zewald JJA, Kerkmeer M, Feitsma M, van Rijssel DA. The impact of teach-back on patient recall and understanding of discharge information in the emergency department: the Emergency Teach-Back (EM-TeBa) study. Int J Emerg Med. 2020 Sep 24;13(1):49. doi: 10.1186/s12245-020-00306-9. PMID: 32972361; PMCID: PMC7513274.

4. Levine DM, Linder JA, Landon BE. Characteristics of Americans With Primary Care and Changes Over Time, 2002-2015. JAMA Intern Med. 2020 Mar 1;180(3):463–466. doi: 10.1001/jamainternmed.2019.6282. PMID: 31841583; PMCID: PMC6990950.

5. IHS Markit, Ltd.: The Complexities of Physician Supply and Demand: Projections From 2019 to 2034. AAMC, Washington, DC; 2021. https://www.aamc.org/data-reports/workforce/data/complexities-physician-supply-and-demand-projections-2019-2034.

6. Kyriacou DN, Handel D, Stein AC, Nelson RR. BRIEF REPORT: Factors affecting outpatient follow-up compliance of emergency department patients. J Gen Intern Med. 2005 Oct;20(10):938–42. doi: 10.1111/j.1525-1497.2005.0216_1.x. PMID: 16191142; PMCID: PMC1490224.

7. Paterick TE, Patel N, Tajik AJ, Chandrasekaran K. Improving health outcomes through patient education and partnerships with patients. Proc (Bayl Univ Med Cent). 2017 Jan;30(1):112–113. doi: 10.1080/08998280.2017.11929552. PMID: 28152110; PMCID: PMC5242136.

8. Thomas EJ, Burstin HR, O’Neil AC, Orav EJ, Brennan TA. Patient noncompliance with medical advice after the emergency department visit. Ann Emerg Med. 1996 Jan;27(1):49–55. doi: 10.1016/s0196-0644(96)70296-2. PMID: 8572448.

9. Iyeke LO, Razack B, Richman M, Berman AJ, Davis F, Willis H, Gizzi-Murphy M, Guilherme S, Johnson S, Njoku C, Ramjattan G, Krol K, Kwon N. Novel Discharge Center for Transition of Care in Vulnerable Emergency Department Treat and Release Patients. Cureus. 2023 Feb 13;15(2):e34937. doi: 10.7759/cureus.34937. PMID: 36938288; PMCID: PMC10017056.

10. Learning organization - Wikipedia. Available at: https://en.wikipedia.org/wiki/Learning_organization. Accessed 1/27/24.

11. Schrader CD, Robinson RD, Blair S, Shaikh S, d’Etienne JP, Kirby JJ, Cheeti R, Zenarosa NR, Wang H. Identifying diverse concepts of discharge failure patients at emergency department in the USA: a large-scale retrospective observational study. BMJ Open. 2019 Jun 27;9(6):e028051. doi: 10.1136/bmjopen-2018-028051. PMID: 31248927; PMCID: PMC6597618.

